# Cultural engagement and mental disorders: A prospective negative control analysis of the English Longitudinal Study of Ageing with linked Hospital Episode Statistics

**DOI:** 10.64898/2026.06.05.26354991

**Authors:** Pei Qin, Andrew Steptoe, Daisy Fancourt

## Abstract

Cultural engagement is associated longitudinally with better mental health and reduced depression incidence, but evidence has largely relied on self-reported symptoms and diagnoses, leaving uncertainty about clinically recorded disorders, and residual confounding remains a concern. Here, we examined whether cultural engagement (including going to cinemas, museums, galleries, exhibitions, theatre, concerts, or opera) predicts hospital-treated mental disorders in 8,274 adults aged 50 years or older from the English Longitudinal Study of Ageing. Participant records were linked to ICD-10 diagnoses in Hospital Episode Statistics and mortality records with follow-up of up to 20 years. In fully adjusted Cox models accounting for sociodemographic, lifestyle, and social factors and multiple testing, frequent cultural engagement was associated with lower risk of any mental disorders (HR 0.71, 95% CI 0.62-0.82, P_FDR_<0.001), dementia (0.71, 0.56-0.89, P_FDR_=0.010), substance misuse (0.75, 0.59-0.95, P_FDR_=0.040), and mood disorders (0.73, 0.56-0.95, P_FDR_=0.044), but not neurotic disorders. Associations persisted after excluding early incident cases and adjusting for baseline depressive symptoms and cognition, and showed robustness to unmeasured confounders. To further probe causality, eye disease, ear disease, and traumatic brain injury – which share similar socio-demographic profiles to mental disorders - were prespecified as negative control outcomes. Cultural engagement was not associated with any negative control outcomes. These findings provide triangulated statistical data to suggest that cultural engagement is associated with reduced risk of several clinically recorded mental disorders and support further testing of cultural engagement as a population mental health strategy.

## Introduction

The arts encompass a diverse range of human practices relating to the production or experience of human creativity and imagination. Arts engagement can include actively participating in the arts (e.g., dance, music, crafts; hereafter “arts participation”) and attending cultural events (e.g., going to live music events, museums, galleries, theatre performances, and the cinema; hereafter “cultural engagement”) (1). There is increasing recognition that arts and cultural engagement are health-promoting behaviours that impact a wide range of health outcomes (2–4). Complexity science theories posit that arts and cultural engagement comprises multiple salutogenic ingredients (physical, cognitive, social, creative) that activate diverse mechanistic pathways that causally influence health (5–7). In terms of psychological mechanisms, arts and cultural engagement modulates affect, emotion regulation and self-referential processing, thereby reducing maladaptive rumination and enhancing emotional resilience. Cognitively, engagement activates diverse neurological regions, including those involved in memory and executive functioning, and enhances functional connectivity between brain regions vulnerable to ageing. Biologically, engagement attenuates psychophysiological stress response, including reducing hypothalamic–pituitary–adrenal axis activation and downstream inflammatory processes. Socio-behaviourally, arts and cultural engagement can increase social bonding, prosociality, and reduce loneliness, as well as promoting adaptive routines and reinforce health-supportive habits. Indeed, many of these mechanistic processes parallel those formally integrated within cognitive-behavioural, humanistic, psychodynamic and postmodern psychotherapeutic treatments (8). As such, cultural engagement can be conceptualised as a health-promoting behaviour that presents a distributed, low-intensity therapeutic system embedded in everyday life (4, 8).

Epidemiological research on arts and cultural engagement has most commonly focused on outcomes related to mental health and brain health. Both cross-sectional and longitudinal studies have demonstrated that arts and cultural engagement are associated with better mental wellbeing (9–12), lower levels of self-reported depressive symptoms and anxiety (12–16), and better preservation of cognition (17, 18). Some studies have also reported potential preventative effects on incident mental disorders. For example, one study using data from the English Longitudinal Study of Ageing (ELSA) found a dose–response relationship between frequency of cultural engagement and reduced risk of incident depression in older adults over the following decade, independent of sociodemographic, health-related and social confounders (13). Another longitudinal study in the Danish general population found associations between low arts and cultural engagement and both new onset and persistent depression (16). And a third found reduced incidence of and a longer time to dementia amongst people with more frequent cultural engagement (19). However, there remain a number of gaps in the literature.

First, previous research is limited to self-reported diagnoses of mental disorders, which could be subject to recall or social desirability biases, or to validated symptom scales that may not capture diagnoses that have been made but are now well-managed with medication/psychotherapy. We lack studies that use objective diagnostic data from electronic patient health records. Second, studies on incidence have focused exclusively on depression to date. Both theoretical work positioning arts within a transdiagnostic, process-based model of mental health, and experimental work demonstrating causal effects on a wide range of psychiatric conditions suggest the arts and cultural engagement may have a preventative effect on a broader range of mental disorders (2, 8, 20). Third, arts and cultural engagement are behaviours that typically shows socio-demographic patterning. Previous studies have used a variety of statistical methods to explore independence of effects, including conditioning on a broad range of identified confounders, applying fixed effects methods that consider time-varying and unmeasured confounders, and using instrumental variable approaches. However, confounding remains an important consideration, and triangulation of findings from a variety of different statistical methods is important to strengthen causal certainty. Approaches such as negative control outcomes have yet to be applied to considerations of cultural engagement and mood disorders (21). Finally, there have been concerns that previously-reported associations between arts and cultural engagement and incident mental disorders could be byproducts of prodromal effects, with early symptoms leading to a progressive reduction in leisure activities (22). There is limited longitudinal data extending beyond 10-year follow-up periods, which could help to disentangle reverse causal effects.

To fill in these knowledge gaps, the present study aimed to investigate the association between cultural engagement and hospital-treated mental disorders. To do so, it leveraged nationally-representative data in the English Longitudinal Study of Ageing (ELSA) linked to UK national Hospital Episode Statistics (HES) records of conditions requiring hospital treatment - either admitted care or as an outpatient - in the National Health Service (NHS) Central Registry. We focused on the overall ICD-10 chapter of mental disorders (ICD codes F00–F99) as well as the four most common mental disorders within this ICD-10 chapter, including dementia (F00–F03, G30, G31), substance abuse (F10–F19), mood disorders (F30–F39), and neurotic disorders (F40–F48). First, we examined longitudinal associations between cultural engagement and incidence of mental disorders over a follow-up period of up to 16 years, adjusting for nested confounders. Second, we identified three negative control outcomes that show similar confounder profiles to test the influence of potential latent confounding. Third, we tested the persistence of findings when exploring further methodological issues such as reverse causality, measurement error and residual confounding.

## Methods

### Study Design and Sample

The present analysis was based on ELSA, a nationally representative panel study of participants aged 50 years or over living in private households in England. ELSA was commenced in 2002, with follow-up every two years. In this study, we analysed data at Wave 4 (2008/2009) as our baseline because this wave had the largest number of the measurement of cultural engagement and covariates considered. ELSA received ethical approval from the National Research Ethics Service (07/H0716/48). The details of ELSA have been described in previous reports (23). This study followed the Strengthening the Reporting of Observational Studies in Epidemiology (STROBE) reporting guideline for cohort studies.

Participants were eligible for inclusion in the sample if they had given consent to the data linkage and were aged 50 years and over at baseline and had complete data on cultural engagement at baseline (Figure 1). This resulted in a total of 8,464 individuals included for prospective analysis of all-cause mortality, although the sample size was then allowed to vary for each ICD-10 code to maximise usage of the available data by only excluding those who had corresponding mental health conditions at or before baseline.

**Figure 1.**
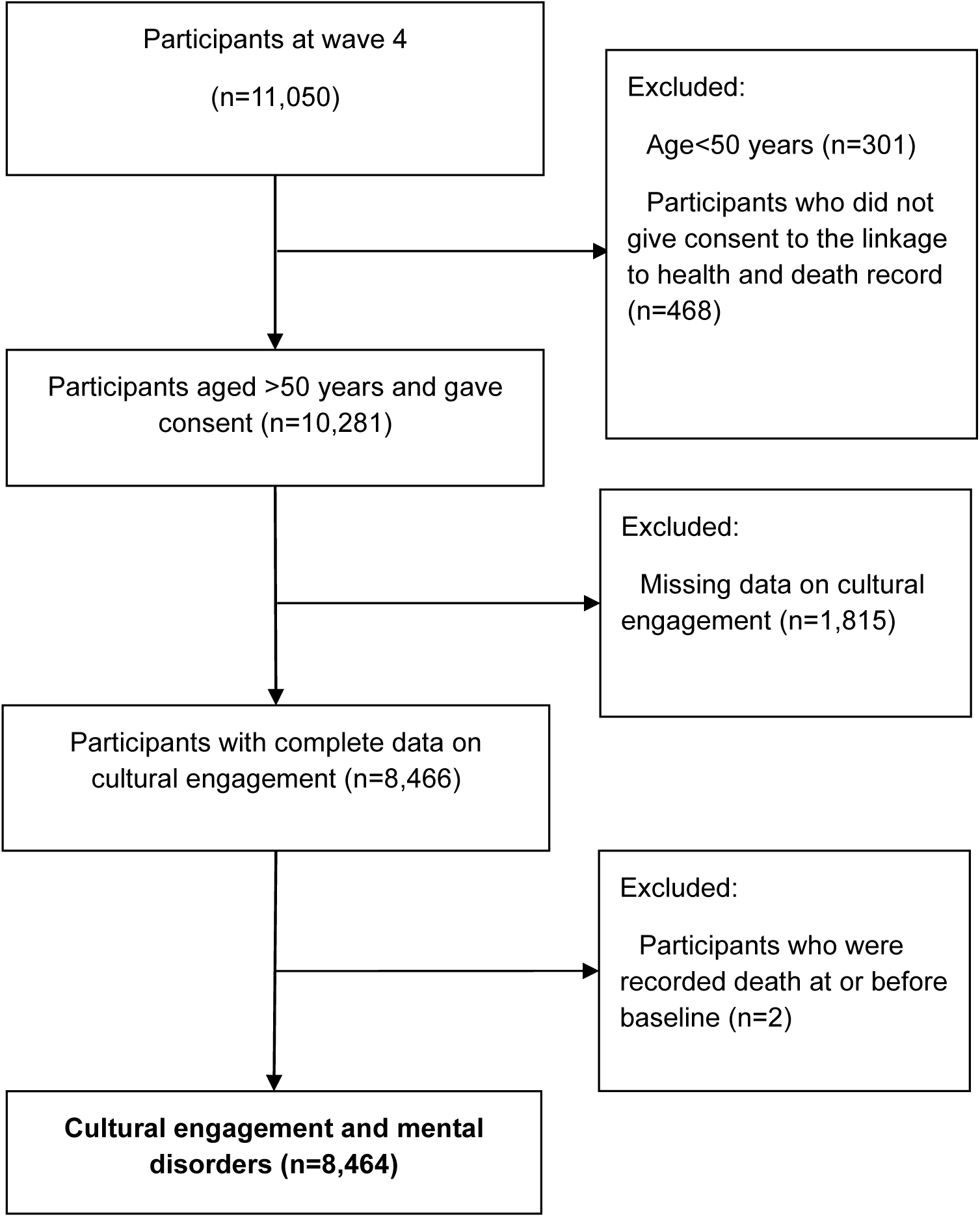
Participants selection.

### Measurement of cultural engagement

We focused specifically on receptive arts activities (i.e., watching or consuming performances or exhibitions). Participants were asked about the frequency of visits to: (a) the cinema, (b) art galleries, exhibitions or museums, (c) theatre, concerts, or opera. Each cultural engagement item was assessed on a five-point scale: 0 (never), 1 (less than once a year), 2 (once or twice a year), 3 (every few months), 4 (once a month), 5 (twice a month or more). Frequency of engagement with any of these activities was categorised as never, infrequent (less than once a year; once or twice a year), or frequent (every few months; monthly or more).

### Measurement of outcomes

ELSA participants who had given consent were linked by unique identification number to the National Health Service’s Central Registry for vital status data up to November 2024, Hospital Episode Statistics (HES) Admitted Patient Care (HES APC) data, and HES Outpatient (HES OP) data up to March 2024. We ascertained mental disorders during follow up on the basis of hospital episode statistics and causes of death, using the International Classification of Diseases Tenth Revision (ICD-10). We focused on the overall mental disorders (F00–F99) and four main mental disorders, including dementia (F00–F03, G30, G31), substance abuse (F10–F19), mood disorders (F30–F39), and neurotic disorders (F40–F48) (Table S1). For our negative controls, we selected eye diseases (H00–H59), ear diseases (H60–H99), and traumatic brain injuries (S2, S6, S71, S78, S79, S97, S98, S99, T04, T06), as these have been demonstrated to have similar demographic and socio-economic patterning to mental disorders in prior publications (24–26). For each health outcome, participants with that specific health condition at or for about 11 years before baseline ascertained from the linked health registry data were excluded from the analysis of incident hospital-treated conditions.

### Covariates

Variables considered likely to confound the associations between cultural engagement and health outcomes were measured at baseline (wave 4). Sociodemographic variables included age (in years), sex (male or female), educational qualifications (university degree or equivalent, including National Vocational Qualifications (NVQ) 4–5; A level/higher education or equivalent including NVQ3; the General Certificate of Education incl. Ordinary level qualification (GCE/O level) or equivalent including NVQ2; other or no educational qualification), ethnicity (White or non-White), total net non-housing wealth (categorised in 5 quintiles), marital status (married/cohabiting or not), employment status (fulltime, part-time, not in employment), and occupational status (managerial and professional occupations, intermediate occupations, small employers and self employed, lower supervisory and technical occupations, and semi-routine occupations; categorised using the five item National Statistics Socio-economic Classification). We used data provided in the nearest subsequent waves if they were missing at baseline.

Lifestyle confounders included smoking status (never, ever and current smokers), frequency of alcohol consumption (>=5 days/week or not), and physical activity level. Physical activity level was categorized into three groups: vigorous (engaging in vigorous activity ≥once a week), moderate (engaging in moderate activity ≥once a week), and inactive (all others). Social covariates included whether living alone (yes or no), being social isolated (yes or no), and being involved with no other community activities (yes or no). Social isolation was measured as whether participants had low contact (less than monthly contact via phone, email or face-to-face) with any social ties (friends, children and wider relatives). Community activities were defined as involvement in any community activities, including being a member of a political party or environmental group, a tenants or neighbourhood watch group, a church or religious association, a charitable association, an education, a social club, a sports, gym or exercise class, or any other society.

In sensitivity analyses we explored further factors that could influence morbidity (particularly if indicative of prodromal effects of imminent morbidity on cultural behaviours) but were less reliable as confounders as they could also lie on the causal pathway. The 8-item version of the Center for Epidemiologic Studies Depression scale (CES-D) was used to measure depression, with the score split into highest quartile vs other quartiles. Memory function was measured by summing scores on immediate and delayed memory (ranging from 0 to 20), while executive function was assessed using an animal naming test (semantic verbal fluency), where participants were asked to list as many animals as possible within one minute. Higher overall scores reflect better memory and executive function, and as for depression the score was split into lowest quartile indicating poorest cognitive functioning vs other quartiles.

### Statistical analyses

The Kaplan-Meier method was used to estimate the cumulative incidence of mental disorders and negative control diseases, and differences in curves were tested by log-rank test. Separate Cox proportional hazard regression models were used to estimate the hazard ratios (HRs) and 95% confidence intervals (CIs). Proportional hazard assumptions were tested using Schoenfeld residuals tests. Variation inflation factors (VIFs) were calculated to test the presence of multicollinearity among the variables. Four models were presented: model 0 adjusted for age and sex; model 1 additionally adjusted for ethnicity, educational qualifications, wealth, marital status, job, and employment; model 2 additionally adjusted for smoking status, alcohol consumption, physical activity level; model 3 additionally adjusted for social factors, including living alone, whether being social isolated, and whether reporting no community activities.

To visualize changes in risk and relative strength of associations across outcomes, Manhattan plots were used to show overall associations between cultural engagement and mental disorders over the follow-up period.

We performed a series of sensitivity analyses to assess potential biases. To examine the extent of unmeasured confounding, E values were calculated. To reduce the influence of prodromal effects altering cultural behaviours, participants who had incident disease within 2 years were excluded, and additionally we ran a further analysis for overall mental disorders additionally adjusting baseline depression and cognition depression and cognition at baseline. To explore potential dose-response relationship reported by our previous study (27) and possible measurement error, frequency of cultural engagement was treated as a continuous variable. (27). To check the robustness of the findings in longer follow-up, wave 2 were treated as baseline, with up to 20 years of follow-up.

All analyses were conducted using R, version 4.3.2 (R Foundation for Statistical Computing). Two-sided p-values were calculated and then corrected for multiple testing by using the Benjamini-Hochberg false discovery rate (FDR) approach (28). Findings were considered statistically significant if the FDR adjusted p-value (P_FDR_) was less than 0.05.

## Results

We analysed data from 8,464 adults over the age of 50 in ELSA who had given consent to data linkage with HES records and completed the self-completion questionnaire in Wave 4 (2008–9) of the study. Table 1 presents the demographics of this analytic sample. Overall, 55.2% were women, and 97.8% were of White ethnicity, and the sample had a mean age of 65.4 years (SD 9.6) at baseline.

**Table 1.** Baseline characteristics of participants overall and by cultural engagement.

|  | Overall | Arts Engagement (%) |  |  | P value |
| --- | --- | --- | --- | --- | --- |
|  |  | Never | Infrequent | Frequent |  |
| N | 8464 | 1566 | 3582 | 3316 |  |
| Age (years) | 65.4 (9.6) | 69.9 (10.9) | 65.0 (9.4) | 63.6 (8.5) | <0.001 |
| Women, n (%) | 4672 (55.2) | 816 (52.1) | 1946 (54.3) | 1910 (57.6) | 0.001 |
| Education group (%) |  |  |  |  | <0.001 |
| No educational qualifications | 2138 (25.3) | 822 (52.7) | 888 (24.9) | 428 (12.9) |  |
| GCE/O-levels/ NVQ 2 | 2715 (32.2) | 464 (29.7) | 1283 (35.9) | 968 (29.2) |  |
| NVQ3/GCE/A-levels | 692 (8.2) | 72 (4.6) | 308 (8.6) | 312 (9.4) |  |
| Higher qualification/NVQ4/NVQ5/degree | 2899 (34.3) | 203 (13.0) | 1094 (30.6) | 1602 (48.4) |  |
| Ethnicity: White (%) | 8278 (97.8) | 1523 (97.3) | 3503 (97.8) | 3252 (98.1) | 0.168 |
| Marital status: married or cohabiting (%) | 5915 (69.9) | 903 (57.7) | 2612 (72.9) | 2400 (72.4) | <0.001 |
| Wealth quintile (%) |  |  |  |  | <0.001 |
| Lowest fifth | 1659 (20.0) | 586 (38.1) | 685 (19.5) | 388 (12.0) |  |
| 2nd fifth | 1658 (20.0) | 406 (26.4) | 761 (21.6) | 491 (15.2) |  |
| 3rd fifth | 1658 (20.0) | 254 (16.5) | 755 (21.4) | 649 (20.1) |  |
| 4th fifth | 1658 (20.0) | 192 (12.5) | 695 (19.7) | 771 (23.9) |  |
| Highest fifth | 1658 (20.0) | 102 (6.6) | 625 (17.8) | 931 (28.8) |  |
| Employment: Working full or part time | 2887 (35.9) | 252 (16.6) | 1260 (36.9) | 1375 (44.3) | <0.001 |
| Occupational status |  |  |  |  | <0.001 |
| Managerial and professional | 2927 (35.7) | 252 (16.7) | 1158 (33.4) | 1517 (46.9) |  |
| Intermediate | 1209 (14.7) | 162 (10.7) | 525 (15.2) | 522 (16.2) |  |
| Small employers and self employed | 951 (11.6) | 167 (11.0) | 416 (12.0) | 368 (11.4) |  |
| Lower supervisory and technical | 737 (9.0) | 198 (13.1) | 351 (10.1) | 188 (5.8) |  |
| Semiroutine | 2383 (29.0) | 734 (48.5) | 1012 (29.2) | 637 (19.7) |  |
| Smoking status (%) |  |  |  |  | <0.001 |
| Never | 3383 (40.1) | 478 (30.6) | 1400 (39.2) | 1505 (45.5) |  |
| Ever | 3969 (47.0) | 718 (45.9) | 1687 (47.2) | 1564 (47.3) |  |
| Current | 1094 (13.0) | 367 (23.5) | 487 (13.6) | 240 (7.3) |  |
| Alcohol: >=5 days/week (%) | 1941 (23.1) | 260 (16.7) | 776 (21.8) | 905 (27.4) |  |
| Sedentary (%) | 645 (7.6) | 277 (17.7) | 241 (6.7) | 127 (3.8) | <0.001 |
| Social isolated (%) | 534 (6.3) | 163 (10.4) | 201 (5.6) | 170 (5.1) | <0.001 |
| No community activities (%) | 2310 (28.3) | 734 (51.2) | 1007 (29.1) | 569 (17.5) | <0.001 |
| Live alone (%) | 1855 (21.9) | 516 (33.0) | 696 (19.4) | 643 (19.4) | <0.001 |

### Who is more culturally engaged?

We defined cultural engagement as frequency of visits to the cinema, art galleries, exhibitions, museums, the theatre, concerts, or the opera. Table 1 presents the distribution of cultural engagement by demographic characteristics. 1,566 (18.5%) reported never engaging in any cultural activities, 3,582 (42.3%) engaged infrequently (less than once a year up to twice a year) and 3,316 (39.2%) engaged frequently (every few months or more). Compared to the never engaged group, those who engaged frequently were more likely to be younger, female, married or cohabiting, work full/part time, be employed as managerial and professional or intermediate area, and have higher education qualifications and wealth. They were also less likely to smoke, drink more than 5 days per week, or be socially isolated, and they were more physically active and engaged in more community activities.

### Is cultural engagement related to incidence of mental disorders?

We restricted our sample to individuals who had no medical record of being diagnosed with each type of mental disorder at baseline or in about 11 years prior. Figure 2 shows Kaplan-Meier curves of the incidence of mental disorders by cultural engagement frequency. In Cox proportional hazard regression models adjusted just for age and sex, cultural engagement was related to a lower hazard of developing all mental disorders (Table 2). When adjusting for socio-demographics, health-related behaviours and social connections this association persisted for overall mental disorders (HR for frequent cultural engagement=0.71, 95% CI 0.62-0.82, P_FDR_<0.001), dementia (0.71, 0.56-0.89, P_FDR_=0.010), substance misuse (0.75, 0.59-0.95, P_FDR_=0.040), and mood disorders (0.73, 0.56-0.95, P_FDR_=0.044). For neurotic disorders, the association was attenuated after considering health-related behaviours. Manhattan plots in Figure 3 show the fully-adjusted models of incidence over the follow-up period for overall mental disorders and the specific diagnostic categories.

**Figure 2.**
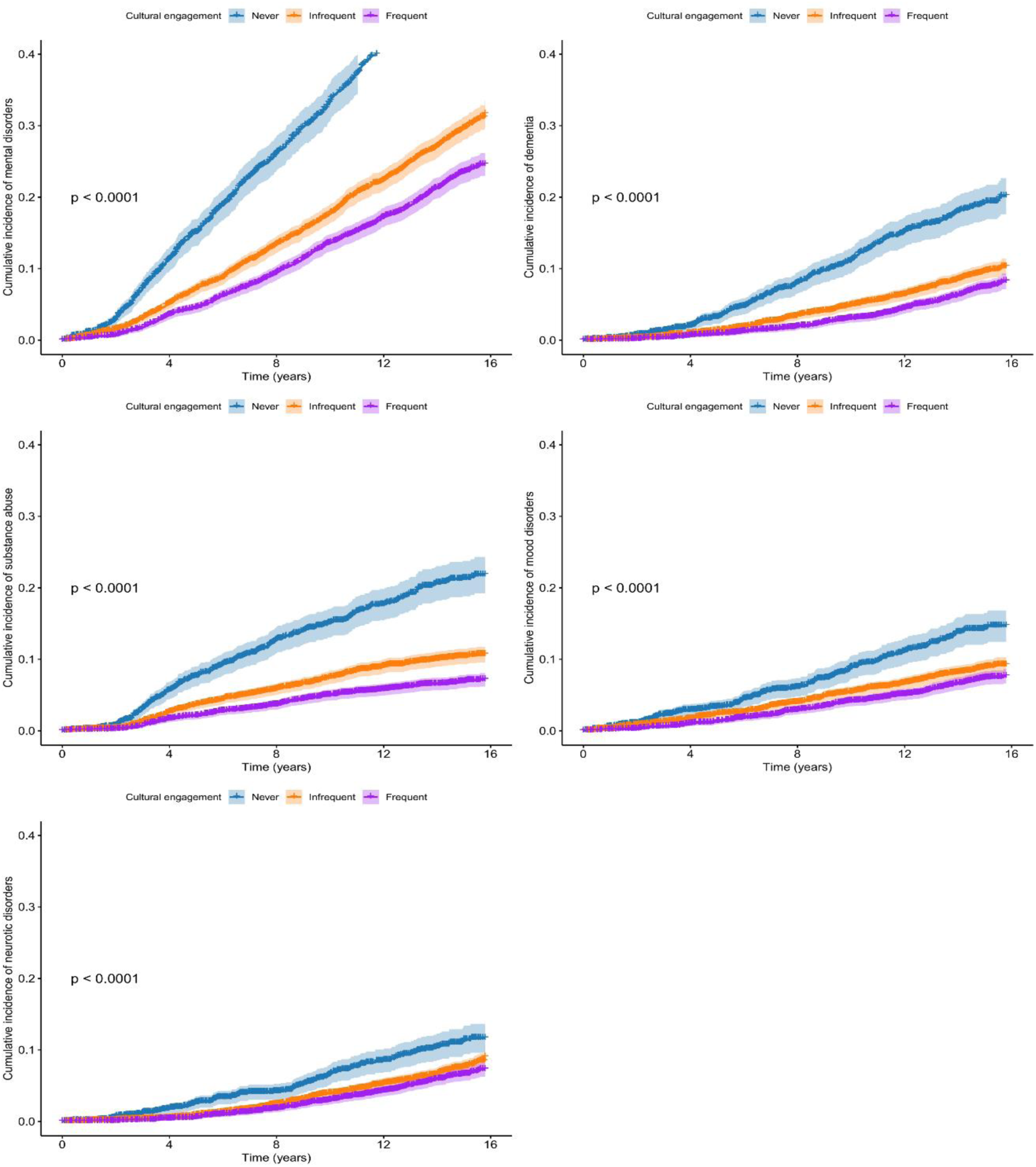
Cumulative incidence for disease categories among people with different cultural engagement groups (never engaged as reference)

**Figure 3.**
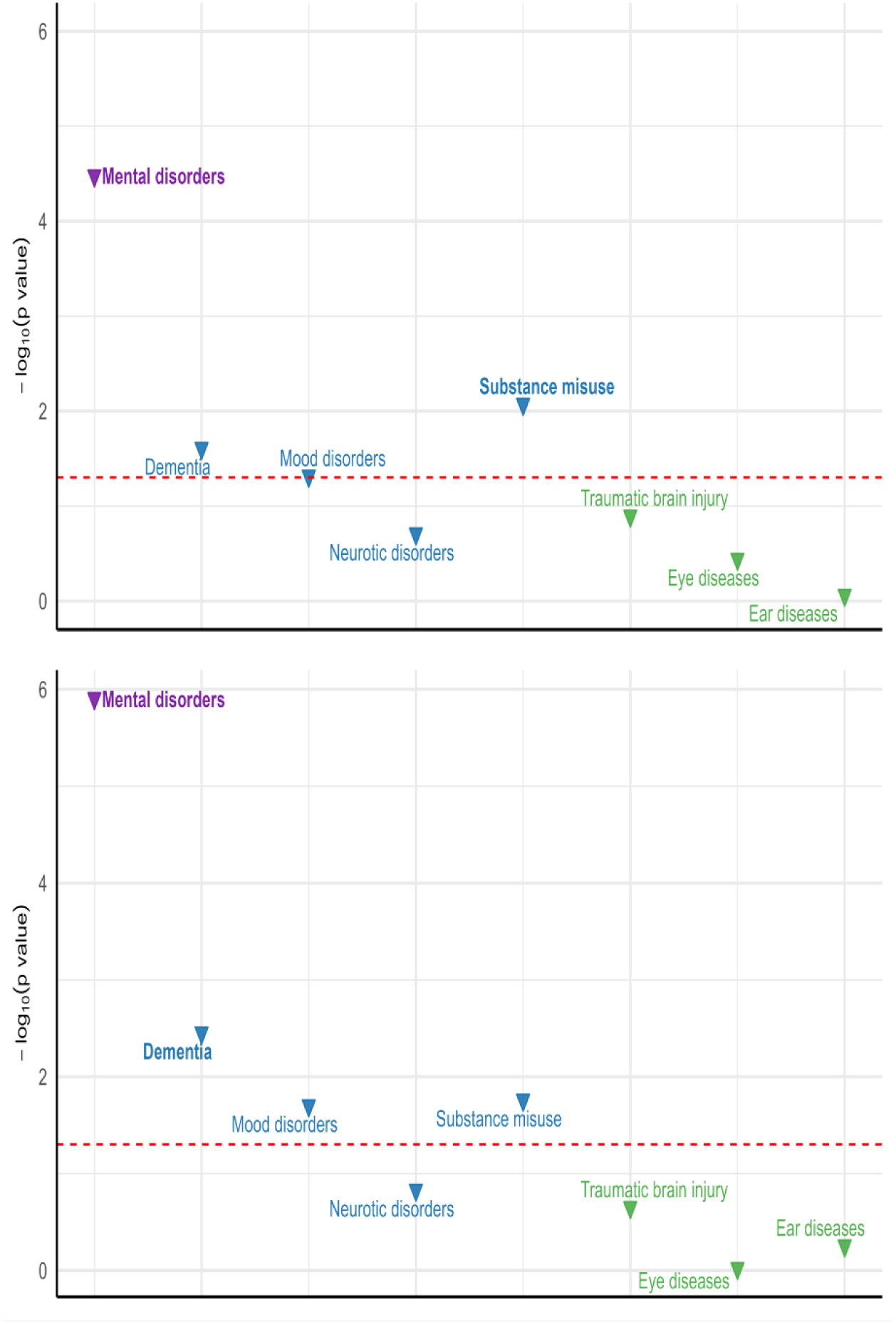
Manhattan plot of the association between cultural engagement and mental disorders. (A) infrequent cultural engagement; (B) frequent cultural engagement. The x-axis represents the disease categorical group, and the y-axis represents the negative log (10) of the P value for the Cox hazard regression model between frequent cultural engagement and disease outcomes. Red horizontal line shows the p value of 0.05, where those above the lone indicates the significant association. Upward and downward triangles indicate hazard ratio (HR) ≥ 1 and HR < 1. Those were labelled bold represent the associations with significance at P < 0.05 (FDR correction for multiple testing). The model was adjusted for age, gender, ethnicity, education, marital status, wealth, employment, and occupational status, lifestyles (smoking, alcohol drinking, sedentary behaviour, and social covariates (living alone, social isolation, and other community activities).

**Table 2.** Association between cultural engagement and mental disorders in the different adjustment models.

| Diseases by category | Model 0 |  |  | Model 1 |  |  | Model 2 |  |  | Model 3 |  |  |
| --- | --- | --- | --- | --- | --- | --- | --- | --- | --- | --- | --- | --- |
|  | HR (95% CI) | P value | FDR adjusted P value | HR (95% CI) | P value | FDR adjusted P value | HR (95% CI) | P value | FDR adjusted P value | HR (95% CI) | P value | FDR adjusted P value |
| Mental disorders |  |  |  |  |  |  |  |  |  |  |  |  |
| Infrequent | 0.61 (0.55-0.67) | <0.001 | <0.001 | 0.72 (0.64-0.81) | <0.001 | <0.001 | 0.78 (0.70-0.88) | <0.001 | <0.001 | 0.77 (0.69-0.87) | <0.001 | <0.001 |
| Frequent | 0.48 (0.43-0.53) | <0.001 | <0.001 | 0.61 (0.54-0.69) | <0.001 | <0.001 | 0.73 (0.64-0.83) | <0.001 | <0.001 | 0.71 (0.62-0.82) | <0.001 | <0.001 |
| Dementia |  |  |  |  |  |  |  |  |  |  |  |  |
| Infrequent | 0.70 (0.58-0.83) | <0.001 | <0.001 | 0.73 (0.60-0.88) | 0.001 | 0.004 | 0.75 (0.62-0.92) | 0.004 | 0.011 | 0.79 (0.65-0.97) | 0.025 | 0.053 |
| Frequent | 0.61 (0.50-0.74) | <0.001 | <0.001 | 0.65 (0.53-0.81) | <0.001 | <0.001 | 0.69 (0.55-0.86) | 0.001 | 0.003 | 0.71 (0.56-0.89) | 0.004 | 0.010 |
| Substance misuse |  |  |  |  |  |  |  |  |  |  |  |  |
| Infrequent | 0.44 (0.37-0.52) | <0.001 | <0.001 | 0.63 (0.52-0.76) | <0.001 | <0.001 | 0.77 (0.64-0.93) | 0.008 | 0.019 | 0.77 (0.63-0.93) | 0.008 | 0.021 |
| Frequent | 0.28 (0.23-0.34) | <0.001 | <0.001 | 0.46 (0.37-0.57) | <0.001 | <0.001 | 0.78 (0.62-0.98) | 0.033 | 0.066 | 0.75 (0.59-0.95) | 0.017 | 0.040 |
| Mood disorders |  |  |  |  |  |  |  |  |  |  |  |  |
| Infrequent | 0.62 (0.51-0.75) | <0.001 | <0.001 | 0.76 (0.61-0.94) | 0.012 | 0.029 | 0.80 (0.64-0.99) | 0.043 | 0.082 | 0.79 (0.63-1.00) | 0.048 | 0.088 |
| Frequent | 0.49 (0.40-0.61) | <0.001 | <0.001 | 0.69 (0.54-0.88) | 0.003 | 0.009 | 0.76 (0.59-0.97) | 0.028 | 0.057 | 0.73 (0.56-0.95) | 0.02 | 0.044 |
| Neurotic disorders |  |  |  |  |  |  |  |  |  |  |  |  |
| Infrequent | 0.66 (0.53-0.82) | **<0.001** | **0.001** | 0.79 (0.62-1.01) | 0.061 | 0.105 | 0.84 (0.65-1.07) | 0.156 | 0.222 | 0.84 (0.65-1.09) | 0.195 | 0.256 |
| Frequent | 0.55 (0.44-0.69) | **<0.001** | **<0.001** | 0.75 (0.58-0.98) | **0.036** | 0.07 | 0.80 (0.61-1.06) | 0.119 | 0.186 | 0.81 (0.61-1.08) | 0.148 | 0.215 |
| **Negative control diseases** |  |  |  |  |  |  |  |  |  |  |  |  |
| **Eye diseases** | 0.91 (0.81-1.03) | 0.131 | 0.195 | 0.95 (0.83-1.07) | 0.394 | 0.442 | 0.94 (0.83-1.07) | 0.324 | 0.384 | 0.94 (0.82-1.07) | 0.362 | 0.422 |
| Infrequent | 0.91 (0.81-1.02) | 0.119 | 0.186 | 1.01 (0.88-1.15) | 0.914 | 0.944 | 1.00 (0.87-1.15) | 0.965 | 0.965 | 1.00 (0.86-1.15) | 0.953 | 0.965 |
| Frequent |  |  |  |  |  |  |  |  |  |  |  |  |
| **Ear diseases** | 0.69 (0.41-1.17) | 0.172 | 0.235 | 0.87 (0.48-1.55) | 0.629 | 0.672 | 0.87 (0.48-1.56) | 0.630 | 0.672 | 0.95 (0.51-1.77) | 0.866 | 0.909 |
| Infrequent | 0.58 (0.33-1.01) | 0.053 | 0.094 | 0.71 (0.37-1.36) | 0.299 | 0.362 | 0.70 (0.36-1.37) | 0.300 | 0.362 | 0.81 (0.40-1.63) | 0.557 | 0.615 |
| Frequent |  |  |  |  |  |  |  |  |  |  |  |  |
| **Traumatic brain injury** |  |  |  |  |  |  |  |  |  |  |  |  |
| Infrequent | 0.82 (0.63-1.08) | 0.164 | 0.229 | 0.77 (0.58-1.03) | 0.077 | 0.13 | 0.78 (0.59-1.05) | 0.102 | 0.168 | 0.79 (0.58-1.07) | 0.127 | 0.194 |
| Frequent | 0.88 (0.67-1.16) | 0.373 | 0.426 | 0.82 (0.60-1.11) | 0.196 | 0.256 | 0.84 (0.62-1.15) | 0.284 | 0.356 | 0.82 (0.59-1.13) | 0.223 | 0.286 |

This table showed the risk estimates compared to the never engaged group.
Model 0 adjusted for age and gender
Model 1 further adjusted for socio-demographic and economic factors (ethnicity, education, marital status, wealth, employment, and occupational status)
Model 2 further adjusted for lifestyle factors (smoking, alcohol drinking, and sedentary behaviour).
Model 3 further adjusted for social covariates (living alone, social isolation and engagement in other community activities).

### Could results be explained by persistent latent socio-economic confounding?

Negative control designs can be used in observational studies to elucidate whether an association is likely to be causal or whether it is a result of unmeasured or residual confounding (29). We selected eye and ear diseases and traumatic brain injuries as the negative control exposure for mental outcomes, because these diseases have similar social patterning to mental disorders reported in the literature, but are not plausibly influenced by cultural engagement. If an association with cultural engagement is observed, it suggests a common confounding structure by shared environment.

Across the follow-up period (max 15.8 years), incidences of overall mental disorders were 28.14%, similar to the 29.51% reported for eye diseases (Table S2). Within each specific diagnosis, incidences ranged from 6.94 to 9.73% for mental disorders, similar to the 4.61% incidence rate for traumatic brain injuries, and a little higher than the 1.09% reported for ear diseases. The outcomes of mental disorders and the negative control outcomes had similar patterns of predictors (Figure 4). For example, mood disorders and neurotic disorders were more common amongst women, as were eye diseases; there was a wealth gradient present strongly across mood disorders, neurotic disorders and eye diseases, and moderately across dementia, substance misuse and ear diseases; and there was no difference based on ethnicity, living alone or social isolation amongst the outcomes or negative controls. Thus, the negative controls were deemed appropriate comparison outcomes.

**Figure 4.**
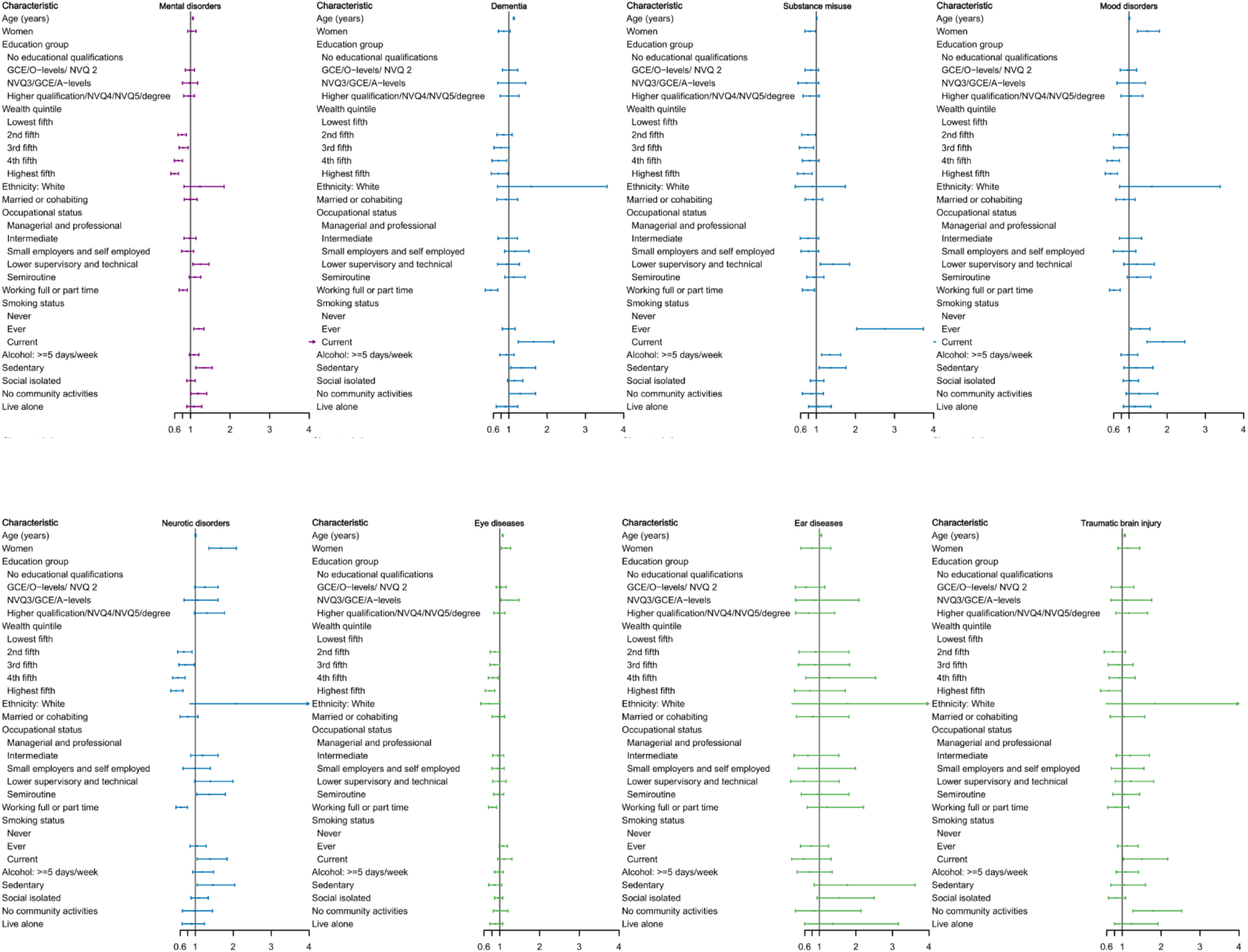
A panel of regression of baseline characteristics predicting each outcome. Cultural engagement and mental disorders: A prospective negative control analysis of the English Longitudinal Study of Ageing with linked Hospital Episode Statistics

We leveraged the negative control outcomes and looked for similar associations with cultural engagement. No associations were observed for the negative control outcomes in the models just adjusted for age and sex, nor across any of the confounding models (Table 2; Figure 3).

### To what extent is the association explained by reverse causality or measurement error?

To evaluate the robustness of our findings, we conducted a series of sensitivity analyses. To test whether results were heavily influenced by reverse causality, such as people with imminent diagnoses already experiencing prodromal symptoms that could influence their cultural engagement, we undertook two further sets of analyses. First, we excluded those with diagnoses shortly after baseline. The results showed that cultural engagement remained significantly associated with mental disorders, dementia, substance misuse and mood disorders in the full adjustment model (Table S3). Second, we additionally adjusted the model for overall mental disorders for baseline depressive symptoms and cognitive functioning. The association remained significant (Table S4).

To consider whether the index that we had created of cultural engagement may be biased to mis-represent patterns of cultural behaviour, we calculated the index in an alternative way that simultaneously considered both frequency and diversity of engagement. The results were broadly aligned, although there was a less evident dose-response relationship for dementia (Table S5).

### How robust are the findings to further unmeasured confounding and longer follow up?

Although our negative control design provides one way of considering residual confounding, it remains possible that further unmeasured or unidentified variables that influence both mental disorders and our negative controls could be acting as confounders. To assess the robustness of the observed association to unmeasured confounding, we calculated E-values, which quantify the minimum strength of association that an unmeasured confounder would need with both the exposure and the outcome to fully explain away an observed association, conditional on the measured covariates (30). The E-values for the point estimates for frequent cultural engagement ranged from 1.77 to 2.17 (Table S6). For comparison, in the overall mental disorder outcome, the strongest measured confounder (smoking status) was associated with the outcome with a risk ratio of 4.62 (95% CI 4.04-5.27) for current smoking, but this was something of an outlier. The second strongest confounder was sedentary behaviours (RR=1.28, 95% CI 1.09-1.49). So, an additional confounder that exceeds the association observed for every other measured confounder currently included in the model would be needed to explain away the observed association entirely.

To assess the robustness of the associations over time, we performed sensitivity analyses by extending the follow-up period up to 20 years, which used wave 2 (2004/2005) as baseline but had lower sample size (7,427). The association remained significantly associated with mental disorders, substance misuse in the full adjustment model, but cultural engagement was also found to be associated with neurotic disorders (Table S7).

## Discussion

In this nationally representative cohort of older adults linked to Hospital Episode Statistics, cultural engagement was associated with a lower hazard of subsequent hospital-treated mental disorders, with particularly consistent associations for dementia, substance misuse, and mood disorders. These findings provide important advances on previous literature reporting preventative effects of arts and cultural engagement. First, they move beyond self-reported diagnoses and symptom scales by using objective clinical diagnoses in electronic patient health records. Second, they highlight that cultural engagement may relate to risk across multiple diagnostic groupings. Third, the combination of long follow-up, nested confounder adjustment, negative control outcomes, and sensitivity analyses provides a stronger causal triangulation framework than has typically been applied. Notably, the magnitude of protective associations identified is comparable to other established behavioural determinants of mental health reported previously such as physical activity, where prospective meta-analyses have reported 17-25% lower risk of incident depression amongst more active individuals (31, 32). This further reinforces the potential salience of cultural engagement as a health-promoting behaviour.

The findings presented are theoretically consistent with transdiagnostic models of mental health, which posit that diverse psychiatric conditions arise from perturbations in shared underlying processes, such as affect regulation, cognitive flexibility, stress responsivity, and social integration. Recent theoretical syntheses have argued that arts and cultural engagement operate as a complex exposure through precisely these multilevel, interacting mechanisms (20). From this perspective, the observed cross-diagnostic associations are not incidental but expected, reflecting modulation of upstream transdiagnostic processes (33). The less consistent findings for neurotic disorders (including attenuation in the main analyses after adjustment for health-related behaviours), however, suggests some nuances in the causal mechanisms activated. Neurotic disorders (F40-F48) comprise phobic disorders (including social phobias), obsessive compulsive disorders, post-traumatic stress and adjustment disorders, dissociative disorders and somatoform disorders, amongst others. Notably, many of these neurotic disorders are more strongly driven by specific vulnerabilities and event-contingent triggers, including conditioning experiences, trauma exposure, and disorder-specific cognitive–behavioural mechanisms, while mood disorders tend to be cumulative, socially patterned, and mediated through modifiable processes such as affect regulation and stress exposure (34). This suggests that arts and cultural engagement have greater protective effects for conditions with an etiological structure that is more gradual and cumulative rather than rapid-onset, event-contingent disorders.

Cultural engagement is a socially patterned behaviour, and concerns about residual confounding have been central to the interpretation of previous findings. Methodologically, these findings extend prior work by applying a more rigorous triangulation framework, including utilising negative outcome controls for the first time (21). Negative controls cannot eliminate all residual confounding, but they do add specificity: if cultural engagement were simply a proxy for generalised privilege, health-seeking, or unmeasured environmental advantage, one might expect more diffuse protective associations across socially patterned outcomes. However, the absence of associations with eye disease, ear disease, and traumatic brain injury—conditions with comparable socio-economic gradients but no plausible causal pathways from cultural engagement—provides evidence of specificity that is not easily explained by generalized health selection or structural advantage. This suggests that cultural engagement is unlikely to be merely a proxy for broader socio-economic or behavioural profiles. Additionally, the extended follow-up period and exclusion of early cases add further reassuring evidence to longstanding concerns regarding prodromal bias. While no single method can establish causality in observational research, the convergence of evidence across these approaches strengthens confidence that the observed associations are not solely artefactual.

Several limitations should be considered. First, cultural engagement was self-reported and measured at a single time point, focusing on only a subset of overall cultural behaviours. As a result, it presents an important insight into how certain cultural behaviours relate to mental disorders. However, future work is encouraged that considers broader definitions. In particular, there is a need for work that takes an exposomic approach to examine the impact of changes in engagement patterns over time, and differential risk-reducing effects of sustained and transient behavioural patterns. Second, although extensive confounders were included, residual confounding cannot be entirely excluded, particularly from unmeasured variables such as personality traits or early-life exposures. While E-value analyses suggest that substantial unmeasured confounding would be required to fully explain the findings, and previous fixed effects analyses have indicated that unmeasured time-invariant confounders have little influence on models (35, 36), it is important to continue probing confounding. Third, the sample was predominantly White and older, which may limit generalisability to more diverse populations or younger age groups. Fourth, the use of hospital-treated outcomes provides important objective data on more severe experiences that warranted hospital mental health care. However, only a minority of cases reach hospital, and many remain undiagnosed entirely. Therefore, it remains important to interpret findings alongside studies that leverage symptom-based scales too. Future research is recommended that explores diagnoses within Mental Health Services data sets when data linkages can be undertaken. Finally, while our analyses included exploring the overall ICD-10 chapter for mental disorders, we were only able to focus on the four most common ICD-10 diagnostic codes within this chapter in our more specific analyses due to very small sample sizes for other diagnoses. Research looking at diagnoses including psychosis and schizophrenia is recommended in data sets with larger sample sizes.

These findings have several implications for future research. Conceptually, they support the framing of cultural engagement within process-based and systems-oriented models of mental health, and emphasise the value of taking outcome-wide approaches to consider the impact between cultural engagement and novel diagnostic groups. Empirically, in line with emerging “cultural exposome” models, future iterations of cohort questionnaires are encouraged to incorporate more measures of qualitative dimensions of cultural engagement, including modality, diversity, intensity, and social context, in order to capture its dynamic and cumulative nature and help elucidate underlying ingredients and causal mechanisms (20). Methodologically, the further application of other advanced causal inference techniques is recommended to continue disentangling causal pathways and provide robust data to enable strong triangulation and strengthen causal claims. All of this could lay the groundwork for future experimental and quasi-experimental studies to identify if individual- or population-based approaches to increase cultural engagement could causally reduce the risk of mental disorders.

Overall, this study strengthens the case for cultural engagement as a potentially important population-level determinant of mental health. Its main contribution is not simply to replicate prior associations, but to show that these associations are detectable for clinically recorded hospital outcomes, span multiple psychiatric domains, and remain after a set of methodological tests specifically designed to challenge common non-causal explanations. In that respect, the findings help move the field from descriptive optimism toward a more mature causal epidemiology of arts and mental health.

## Data Availability

ELSA data are available through registration with the UK data service (https://beta.ukdataservice.ac.uk/datacatalogue/series/series?id=200011). The linked HES and mortality data cannot be shared directly due to data sharing agreements with NHS England. But applications for data access can be made via the UK Longitudinal Linkage Collaboration: https://ukllc.ac.uk/apply.

## Acknowledgment

Authors’ contributions

P.Q., A.S., and D.F. designed research; P.Q., A.S., and D.F. performed research; P.Q. analyzed data; P.Q. and D.F. wrote the paper, A.S. and D.F. reviewed and edited the paper. All authors (P.Q., A.S., and D.F.) are responsible for the decision to submit this paper.

