## Supplemental Tables 1-7 for "Cultural engagement and mental disorders: A prospective negative control analysis of the English Longitudinal Study of Ageing with linked Hospital Episode Statistics"

**Supplemental Table 1.** Included ICD-10 chapters and codes .

| Category | Disease | ICD-10 code |
|---|---|---|
| **Mental disorders** | **Mental disorders** | F00–F99 |
| | Dementia | F00–F03, G30, G31 |
| | Substance abuse | F10–F19 |
| | Psychotic disorders | F20–F29 |
| | Mood disorders | F30–F39 |
| | Neurotic disorders | F40–F48 |
| **Negative control diseases** | **Diseases of the eye** | H00–H59 |
| | **Diseases of the ear** | H60–H99 |
| | **Traumatic brain injury** | S2, S6, S71, S78, S79, S97, S98, S99, T04, T06 |

**Supplemental Table 2.** Case numbers by ICD-10 code

| Disease | N | Cases | Follow-up duration Mean (SD) | Incidence (%) | Incidence rate (per 1000 person-year) |
|---|---|---|---|---|---|
| **Mental disorders** | 8274 | 2258 | 11.97 (4.66) | 28.14 | 22.81 |
| Dementia | 8454 | 750 | 12.98 (4.12) | 8.98 | 6.83 |
| Substance misuse | 8396 | 790 | 12.63 (4.37) | 9.73 | 7.45 |
| Mood disorders | 8389 | 655 | 12.78 (4.24) | 8.23 | 6.11 |
| Neurotic disorders | 8416 | 560 | 12.86 (4.13) | 6.94 | 5.17 |
| **Negative control diseases** | | | | | |
| Eye diseases | 7653 | 2259 | 12.95 (4.15) | 29.51 | 25.18 |
| Ear diseases | 8419 | 92 | 13.10 (4.04) | 1.09 | 0.83 |
| Traumatic brain injury | 8367 | 386 | 11.72 (4.77) | 4.61 | 3.56 |

**Supplemental Table 3.** Association between cultural engagement and mental disorders excluding diseases that developed shortly after baseline

| | Excluding diseases that developed in two years | | |
|---|---|---|---|
| | HR (95% CI) | P value | FDR adjusted P value |
| **Mental disorders** | | | |
| Infrequent | 0.77 (0.68–0.87) | **<0.001** | **<0.001** |
| Frequent | 0.72 (0.63–0.83) | **<0.001** | **<0.001** |
| **Dementia** | | | |
| Infrequent | 0.78 (0.63–0.95) | **0.015** | **0.035** |
| Frequent | 0.70 (0.56–0.89) | **0.003** | **0.009** |
| **Substance misuse** | | | |
| Infrequent | 0.77 (0.63–0.95) | **0.012** | **0.029** |
| Frequent | 0.76 (0.60–0.96) | **0.023** | **0.047** |
| **Mood disorders** | | | |
| Infrequent | 0.74 (0.58–0.95) | **0.017** | **0.036** |
| Frequent | 0.73 (0.56–0.96) | **0.023** | **0.047** |
| **Neurotic disorders** | | | |
| Infrequent | 0.89 (0.68–1.17) | 0.417 | 0.485 |
| Frequent | 0.87 (0.65–1.17) | 0.361 | 0.428 |
| **Negative control diseases** | | | |
| **Eye diseases** | | | |
| Infrequent | 0.96 (0.83–1.11) | 0.568 | 0.606 |
| Frequent | 1.03 (0.88–1.20) | 0.709 | 0.71 |
| **Ear diseases** | | | |
| Infrequent | 0.88 (0.46–1.68) | 0.689 | 0.710 |
| Frequent | 0.69 (0.33–1.43) | 0.313 | 0.409 |
| **Traumatic brain injury** | | | |
| Infrequent | 0.71 (0.52–0.98) | 0.035 | 0.065 |
| Frequent | 0.79 (0.56–1.11) | 0.171 | 0.257 |

Model adjusted for age and gender, ethnicity, education, marital status, wealth, employment, occupational status, smoking, alcohol drinking, and sedentary behaviour, living alone, social isolation and engagement in other community activities).

**Supplemental Table 4.** Association between cultural engagement and mental disorders further adjusting for cognition and depression at baseline

| | HR (95% CI) | p-value |
|---|---|---|
| **Cultural engagement** | | |
| Never | ref | |
| Infrequent | 0.83 (0.74-0.94) | **0.003** |
| Frequent | 0.79 (0.68, 0.90) | **<0.001** |

Model adjusted for age, gender, ethnicity, education, marital status, wealth, employment, occupational status, smoking, alcohol drinking, sedentary behaviour, living alone, social isolation and engagement in other community activities, memory (lowest quartiles vs. others) and executive function (lowest quartiles vs. others) and depression (highest quartiles vs. others) at baseline.

**Supplemental Table 5.** Association between cultural engagement and mental disorders using tertiles of cultural engagement (range 0-15)

| | Full adjustment model (tertile 1 as reference group) | | |
|---|---|---|---|
| **Diseases by category** | **HR (95% CI)** | **P value** | **FDR adjusted P value** |
| **Mental disorders** | | | |
| Tertile 2 | 0.73 (0.65–0.82) | **<0.001** | **<0.001** |
| Tertile 3 | 0.73 (0.64–0.83) | **<0.001** | **<0.001** |
| **Dementia** | | | |
| Tertile 2 | 0.76 (0.62–0.93) | **0.007** | **0.017** |
| Tertile 3 | 0.82 (0.66–1.03) | 0.090 | 0.134 |
| **Substance misuse** | | | |
| Tertile 2 | 0.76 (0.63–0.92) | **0.005** | **0.013** |
| Tertile 3 | 0.73 (0.58–0.93) | **0.011** | **0.023** |
| **Mood disorders** | | | |
| Tertile 2 | 0.72 (0.58–0.89) | **0.003** | **0.007** |
| Tertile 3 | 0.73 (0.57–0.93) | **0.010** | **0.023** |
| **Neurotic disorders** | | | |
| Tertile 2 | 0.81 (0.64–1.02) | 0.070 | 0.112 |
| Tertile 3 | 0.79 (0.61–1.03) | 0.079 | 0.121 |
| **Negative control diseases** | | | |
| **Eye diseases** | 0.93 (0.83–1.05) | | |
| Tertile 2 | 1.06 (0.94–1.21) | 0.237 | 0.31 |
| Tertile 3 | | 0.341 | 0.404 |
| **Ear diseases** | 0.85 (0.47–1.53) | | |
| Tertile 2 | 0.90 (0.47–1.72) | 0.591 | 0.61 |
| Tertile 3 | | 0.749 | 0.749 |
| **Traumatic brain injury** | | | |
| Tertile 2 | 0.80 (0.60–1.06) | 0.12 | 0.164 |
| Tertile 3 | 0.89 (0.66–1.21) | 0.462 | 0.518 |

Model adjusted for age, gender, ethnicity, education, marital status, wealth, employment, occupational status, smoking, alcohol drinking, sedentary behaviour, living alone, social isolation and engagement in other community activities.

**Supplemental Table 6.** E value to consider the unmeasured confounding effect

| Diseases | E-value point estimate (CI) | | | |
|---|---|---|---|---|
| | Model 0 | Model 1 | Model 2 | Model 3 |
| **Mental disorders** | | | | |
| Infrequent | 2.16 (1.97) | 1.82 (1.58) | 1.66 (1.41) | 1.69 (1.44) |
| Frequent | 2.70 (2.47) | 2.16 (1.91) | 1.79 (1.53) | 1.85 (1.56) |
| **Dementia** | | | | |
| Infrequent | 2.21 (1.70) | 2.08 (1.53) | 2.00 (1.39) | 1.85 (1.21) |
| Frequent | 2.66 (2.04) | 2.45 (1.77) | 2.26 (1.60) | 2.17 (1.50) |
| **Substance misuse** | | | | |
| Infrequent | 3.97 (3.52) | 2.55 (1.96) | 1.92 (1.36) | 1.92 (1.36) |
| Frequent | 6.60 (5.33) | 3.77 (2.90) | 1.88 (1.16) | 2.00 (1.28) |
| **Mood disorders** | | | | |
| Infrequent | 2.61 (2.00) | 1.96 (1.32) | 1.81 (1.11) | 1.85 (1.00) |
| Frequent | 3.50 (2.66) | 2.26 (1.53) | 1.96 (1.21) | 2.08 (1.29) |
| **Neurotic disorders** | | | | |
| Infrequent | 2.40 (1.74) | 1.85 (1.00) | 1.67 (1.00) | 1.67 (1.00) |
| Frequent | 3.04 (2.26) | 2.00 (1.16) | 1.81 (1.00) | 1.77 (1.00) |

This Supplemental Table howed the E value of the risk estimates compared to the never engaged group.
Model 0 adjusted for age and gender.
Model 1 further adjusted for socio-demographic and economic factors (ethnicity, education, marital status, wealth, employment, and occupational status).
Model 2 further adjusted for lifestyle factors (smoking, alcohol drinking, and sedentary behaviour).
Model 3 further adjusted for social covariates (living alone, social isolation and engagement in other community activities).

**Supplemental Table 7.** Association between cultural engagement and mental disorders when treating wave 2 as the baseline

| | Full adjustment model (never engaged as reference group) | | |
|---|---|---|---|
| **Diseases by category** | **HR (95% CI)** | **P value** | **FDR adjusted P value** |
| **Mental disorders** | | | |
| Infrequent | 0.77 (0.67–0.87) | **<0.001** | **<0.001** |
| Frequent | 0.70 (0.60–0.81) | **<0.001** | **<0.001** |
| **Dementia** | | | |
| Infrequent | 0.85 (0.69–1.04) | 0.104 | 0.196 |
| Frequent | 0.77 (0.62–0.97) | **0.027** | 0.063 |
| **Substance misuse** | | | |
| Infrequent | 0.61 (0.48–0.77) | **<0.001** | **<0.001** |
| Frequent | 0.66 (0.51–0.86) | **0.002** | **0.007** |
| **Mood disorders** | | | |
| Infrequent | 0.98 (0.75–1.27) | 0.853 | 0.88 |
| Frequent | 0.88 (0.66–1.18) | 0.386 | 0.494 |
| **Neurotic disorders** | | | |
| Infrequent | 0.72 (0.54–0.96) | **0.023** | 0.055 |
| Frequent | 0.58 (0.42–0.80) | **0.001** | **0.003** |
| **Negative control diseases** | | | |
| **Eye diseases** | | | |
| Infrequent | 0.91 (0.80–1.04) | 0.155 | 0.283 |
| Frequent | 0.91 (0.79–1.04) | 0.172 | 0.307 |
| **Ear diseases** | | | |
| Infrequent | 1.02 (0.51–2.03) | 0.959 | 0.97 |
| Frequent | 0.91 (0.43–1.92) | 0.798 | 0.852 |
| **Traumatic brain injury** | | | |
| Infrequent | 1.06 (0.76–1.49) | 0.734 | 0.82 |
| Frequent | 0.94 (0.65–1.36) | 0.749 | 0.82 |

Model adjusted for age, gender, ethnicity, education, marital status, wealth, employment, occupational status, smoking, alcohol drinking, sedentary behaviour, living alone, social isolation and engagement in other community activities.
